# Geometric brain signatures of Alzheimer’s disease progression and subtypes

**DOI:** 10.64898/2026.05.14.26353211

**Authors:** Boning Tong, Trang Cao, Duy Duong-Tran, Christos Davatzikos, Paul Thompson, Saykin Andrew, Alex Fornito, Li Shen, the Alzheimer’s Disease Neuroimaging Initiative

## Abstract

Alzheimer’s disease (AD) patients suffer from consequential diagnostic delay due to the lack of accessible biomarkers. They also show different responses to treatments due to disease heterogeneity and progression. Here, we developed a novel framework to identify disease progression and subtypes by using geometric brain signatures derived from multiple neuroimaging modalities, including [^**18**^F]-Florbetapir (AV45) Positron Emission Tomography (PET), [^**18**^F]-Fludeoxyglucose (FDG) PET, and structural Magnetic Resonance Imaging (MRI). These signatures were derived by decomposing corresponding maps of amyloid-beta levels, metabolic activity, and cortical thickness in terms of the fundamental, resonant modes––eigenmodes––of cortical geometry, each tied to a specific spatial resolution scale. Our results showed that geometric eigenmode-based features identified trajectories of disease progression, quantified as pseudotime, in distinct subtypes. The disease progression trajectories and subtypes are identified with high stability and are highly related to biological and cognitive measures. These performances are superior to those obtained using conventional localised features and remain robust across datasets, indicating that geometric signatures of brain structure and function can be used to uncover new markers of AD diagnosis and prognosis that are missed by conventional localisation approaches.

## 1 Introduction

Alzheimer’s disease (AD) imposes a tremendous burden on patients and carers. Despite recent advances in biomarker identification and treatment, AD patients are still facing delays of months or even years before a diagnosis from the first presentation at primary care. This delay consumes precious time from patients and family for planning disease management and results in poor treatment outcomes and family wellbeing. Even post-diagnosis, it is still not clear how the available drugs affects the disease symptoms and why some drugs are only effective in a subgroup of patients [41, 24]. Therefore, identification of accessible and robust diagnostic and prognostic biomarkers is of paramount importance.

Non-invasive neuroimaging is a promising tool for identifying such biomarkers. Increasingly, the resulting measures, particularly those derived from positron emission tomography (PET) and magnetic resonance imaging (MRI) are being combined with mathematical models to infer trajectories of progression. For example, event-based models estimate a probabilistic sequence of discrete biomarker abnormality events along disease progression [47, 46]; pseudotime inference methods map individuals along a continuous latent trajectory to capture disease progression [60, 21, 35]; and subtype-aware progression models extend event-based models by jointly inferring multiple distinct biomarker progression patterns and assigning individuals to subtypes [58, 61]. While these models have advanced our understanding of disease dynamics, many of them rely on single-modality or population-level features, limiting their ability to capture diverse pathological changes [4]. On the other hand, emerging evidence suggests that integrating complementary data sources through multimodal approaches can substantially improve robustness and sensitivity to early disease-related changes [21, 14, 56, 55, 57].

Building on these ideas, the recently developed multi-omics contrastive trajectory inference (mcTI) algorithm [22] has leveraged complementary molecular modalities to jointly infer continuous disease trajectories and assign individuals to subtypes, providing finer-grained insight into heterogeneous disease patterns. However, the initial application relied on postmortem brain and in vivo blood molecular data; the former limits real-time prognosis, while the latter lacks the anatomical precision to map spatial neurodegeneration. Here, we propose the use of multiple neuroimaging modalities that can be easily accessed through routine clinical assessments for putative subtyping and prognostication.

A key challenge in this work concerns the appropriate neuroimaging phenotypes for inclusion in the model. A vast literature of neuroimaging studies in AD have identified a diverse array of changes across multiple brain regions, defined either at the level of regions-of-interest (ROIs) or image voxels/vertices [10, 18, 33, 32]. These approaches generally assume that these changes (1) can be localised to specific areas; (2) arise independently of each other; and (3) are expressed at a spatial scale defined by the measurement resolution (e.g., a ROI or voxel). They also offer no insights into the fundamental constraints that may have shaped those changes. Recent work [7, 39] has shown that geometric eigenmodes of the brain can be used to derive robust descriptions of diverse aspects of brain anatomy and activity, including the changes observed in AD, while offering insights into the generative physical mechanisms that shape the observed spatial patterns. These modes can be used to describe the natural, resonant modes of brain anatomy and dynamics, thus corresponding to preferred ways in which the brain can respond to some kind of excitation by virtue of its anatomical composition, must like the eigenmodes of a string correspond to its preferred vibrational patterns (also known as string harmonics). Critically, the modes represent an orthogonal basis set that can be used to decompose any spatial brain map, under the assumption that the pattern defined by the map arises from the preferential excitation of distinct, fundamental modes. The modes are ordered by their spatial frequency, offering a spectral decomposition of the data that provides insights into its multiscale organisation, much like a classical Fourier decomposition of a 1-dimensional signal. The geometric eigenmode-based approach has been used to gain insights into brain activity [39], asymmetry [8], connectivity [38], anatomy and disease [7].

Building on these advances, we developed a multimodal trajectory inference framework based on eigenmode-based imaging features and the mcTI algorithm [22]. Specifically, we used geometric eigenmodes to decompose spatial maps of cortical beta-amyloid deposition derived from [^18^F]-Florbetapir (AV45) PET, glucose metabolism from [^18^F]-Fludeoxyglucose (FDG) PET, and cortical thickness from structure MRI (sMRI), and used the resulting geometric signatures as inputs to the mcTI model to identify disease progression score, defined as pseudotime, and assign AD sub-types based on pseudotime with cross-validation. We evaluated the validity of the inferred disease progression pseudotime estimates and putative subtypes by their distinct associations with biological and cognitive measures. Our results showed that eigenmode-based features can be used to identify putative AD subtypes with distinct pseudotime illness trajectories that are more stable with permutation testing and robust across datasets than features extracted using conventional approaches. These results highlighted the potential of our proposed framework for AD diagnosis and prognosis and demonstrate that the potential value of geometric brain signatures for understanding AD.

## 2 Results

We extracted 30-dimensional mode-based features per hemisphere from person-specific brain maps of amyloid-beta (A*β*) (from AV45 PET), metabolic activity (from FDG PET), and cortical thickness (from sMRI) in the Alzheimer’s Disease Neuroimaging Initiative (ADNI) cohort [51] (Fig. 1 and Methods 4.1). The mode-based features were obtained by decomposing each brain map as a linear combination of eigenmodes of cortical geometry. The eigenmodes were derived using the Laplace-Beltrami Operator on the FsAverage cortical mesh (Methods 4.1.2). The resulting spectrum of coefficient weights from the decomposition reflects the contribution of each mode to the observed maps (Fig. 1(ii.a)). Note that the modes are ordered from low-frequency long-wavelength to high-frequency short-wavelength patterns (Fig. S1). The number of modes (i.e., 30) used for extracting features was optimised using the Spearman correlation between the estimated pseudotime and clinical diagnosis (Methods 4.3).

**Fig 1.**
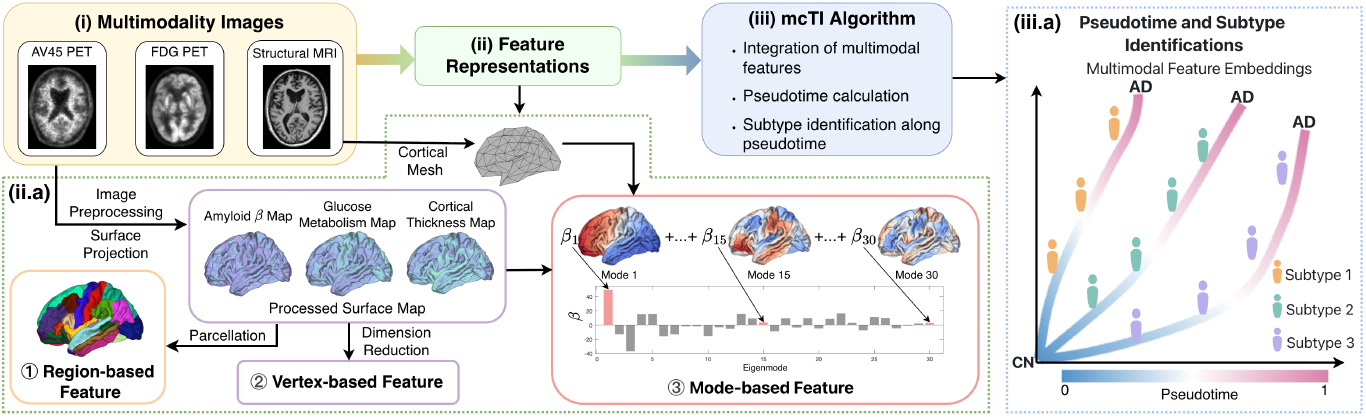
Multimodal imaging pseudotime and subtype identification framework. The framework includes three stages: (i) acquisition of multimodal imaging scans, (ii) feature extraction, and (iii) pseudotime/subtype inference using the mcTI algorithm. (ii.a) Voxel-based AV45 PET, FDG PET, and sMRI images were processed to extract region-based (68 DK atlas cortical regions), vertex-based (PCA-reduced surface maps), and mode-based features. Mode-based features (shown in red box) were obtained by decomposing vertex-based maps onto eigenmodes derived from the cortical mesh. The coefficients (*β*, shown as a bar plot) representing each mode’s contribution and are used as the final mode-based features. (iii.a) Each feature type was independently input into mcTI to compute pseudotime and assign subtypes. Multimodal features were then fused to generate integrated embeddings. As pseudotime increases, subjects progress from cognitively normal (CN) to Alzheimer’s disease (AD) along distinct trajectories. Divergence of these trajectories reflects distinct disease subtypes (three shown as examples), each capturing a coherent progression pattern.

We then applied the mcTI algorithm (Fig. 1(iii.a) and Methods 4.2) to the multimodal mode-based features in order to obtain (i) a pseudotime representing the disease progression score for each participant and (ii) subtypes of particpants with distinct disease progression trajectories. The pseudotime was normalized to the range [0,1], reflecting the relative position of each subject along the inferred AD progression trajectory, with lower values corresponding to cognitive normal (CN) and higher values corresponding to AD at the later-stage. The mcTI algorithm first calculates the pseudotime of a participant as the shortest distance from that participant to the centroid of CN group in the multimodal fused network (FN) generated from multimodal imaging features (Methods 4.2). The mcTI algorithm then randomly assigns participants to subtypes and uses the multimodal features of participants in each subtype to predict the pseudotime of a leave-one-out participant in that subtype. The mcTI algorithm decides the subtypes with the minimum prediction errors and validates their stability with permutation test (Methods 4.4.4). Each subtype identified by this method comprises participants that have similar multimodal features defining the distinct disease progression trajectory. In the following, we show the results of (i) the pseudotime calculation and validation, (ii) the subtype identification and characterisation, and (iii) comparison across different imaging feature extractions.

### 2.1 Identifying and characterising pseudotime as a disease progression score

We first visualise how the predicted pseudotime is distributed in the feature representation space by performing multidimensional scaling (MDS). The MDS embeddings show the pseudotime identified by our framework (Fig. 2a) in comparison with clinical diagnosis (cognitive normal-CN, mild cognitive decline-MCI, and Alzheimer’s disease-AD) given by the dataset (Fig. 2b), where each point represents a participant. The embedding reveals a continuous trajectory ranging from CN to AD. Notably, the MDS space reveals a fan-shaped progression pattern: subjects at lower pseudotime exhibit high dispersion, reflecting the diverse biological signatures of healthy aging and early-stage impairment. Conversely, as pseudotime increases, subjects converge toward a more compact and homogeneous phenotypic state. This convergence suggests that while the initial stages of AD are characterized by significant individual heterogeneity, the late-stage disease process follows a more stereotyped and biologically constrained pathological terminal.

**Fig 2.**
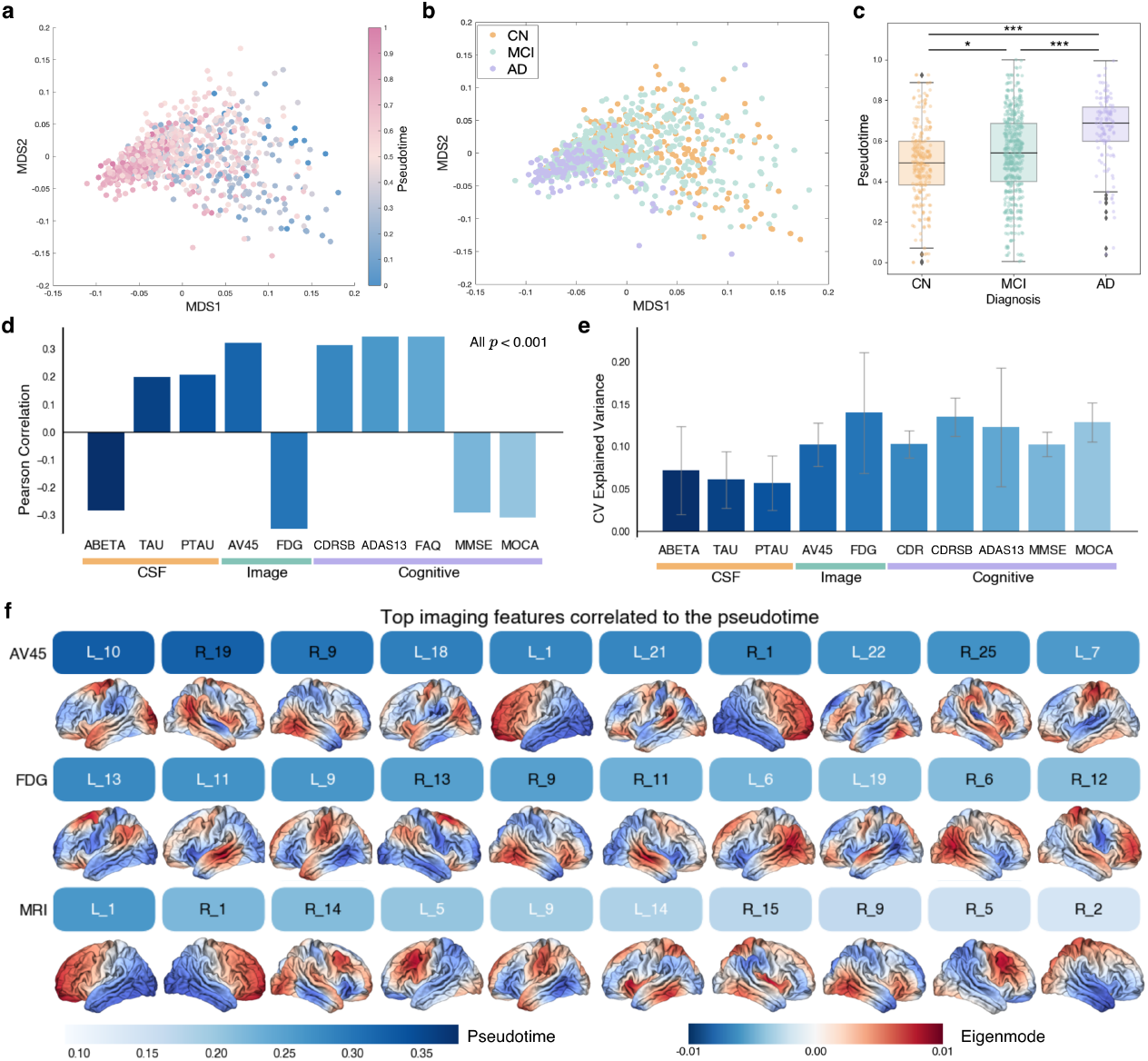
Identifying and characterising pseudotime as a disease progression score. (a–b) Two-dimensional embedding of the fused multimodal imaging feature network obtained using MDS, where each point represents a participant. (a) Points are colored by pseudotime values, where values closer to 1 indicate later stages of AD. (b) Points are colored according to the participant’s clinical diagnosis. (c) The distribution of pseudotime in each clinical diagnosis. Asterisks denote two-sided permutation-based p-values (Bonferroni corrected, ***: *p* < 0.001, **: *p* < 0.01, *: *p* < 0.05). (d) Pearson correlation between pseudotime and different biomarkers, the Bonferroni corrected p-values of all biomarkers are less than 0.001. (e) Mean explained variance values of 5-fold cross-validation regression, using pseudotime to predict each individual biomarker. Error bar indicates the standard deviation.(f) Top ten imaging features ranked by their correlation with pseudotime for each imaging modality. The blue heatmap encodes the magnitude of the correlations, with darker colors indicating stronger associations. Feature identities are annotated, where L and R denote the left and right hemispheres, and indices 1–30 correspond to the 30 eigenmodes. Brain visualisations for each eigenmode are displayed below the corresponding heatmap patches.

We then characterise the clinical, biological, and cognitive relevance of the inferred pseudotime, in line with prior work in the literature[1, 2, 42, 44]. Fig. 2c shows that pseudotime increased monotonically across clinical diagnostic groups (*p* < 0.001; Methods 4.4.1), successfully capturing the continuum of cognitive decline. To evaluate the biological relevance of pseudotime, we calculated its Pearson correlations (Fig. 2d; Methods 4.4.2) with established biomarkers across three distinct domains: (i) cerebrospinal fluid (CSF) measures, including amyloid *β* (ABETA), total tau (TAU), and phosphorylated tau (PTAU); (ii) measures from imaging scans, including AV45 PET and FDG PET summary standardized uptake value ratios (SUVR) [3]; and (iii) clinical cognitive scales, including the Clinical Dementia Rating Sum of Boxes (CDRSB), Alzheimer’s Disease Assessment Scale–Cognitive Subscale 13 (ADAS13), Functional Activities Questionnaire (FAQ), Mini-Mental State Examination (MMSE), and Montreal Cognitive Assessment (MOCA). Higher pseudotime was significantly associated with increased CSF TAU, PTAU, and AV45 summary SUVR, as well as decreased CSF ABETA and FDG summary SUVR, in agreement with known biological patterns of disease progression [18, 33, 32]. Higher pseudotimes were associated with higher CDRSB, ADAS13, and FAQ scores, as well as lower MMSE and MOCA scores. The direction of these correlations is consistent with the known relationships between these cognitive measures and AD progression [50]. To further evaluate the generalizability of these relationships in unseen data, we used 5-fold cross-validated regression to assess whether the derived pseudotime can predict biomarker measurements and reported the mean explained variance in Fig. 2e (Methods 4.4.3). Aligned with the Pearson correlation results, imaging biomarkers showed relatively higher explained variance, while cognitive measures such as ADAS13 and MOCA demonstrated comparable contributions. In contrast, CSF biomarkers exhibited weaker associations. Overall, these results suggest that the inferred pseudotime primarily captures neurodegenerative and clinical progression signals.

We investigated the impacts of the input mode-based features to the resulting pseudotime by quantifying Pearson correlation between them. Fig. 2f highlights the ten most strongly associated modes for each modality along with their corresponding correlation demonstrated in blue scale. Across modalities, AV45-derived features showed the strongest correlations with pseudotime, suggesting that amyloid burden serves as a robust proxy for the inferred disease progression axis. In contrast,MRI-based features exhibited comparatively weaker associations, possibly reflecting greater inter-individual variability of structural atrophy [58]. The strongly-associated modes overlap regions previously implicated in AD in the literature such as the precuneus, posterior cingulate cortex, medial temporal regions, and lateral temporoparietal cortex [9, 43, 53]. However, these modes are predominantly low frequency, suggested that brain changes in AD happen at large-scale or whole-brain levels, contrasting with the focus on finding specific affected regions in the literature [10, 18, 33, 32]. This shift from “focal lesion” to “system-wide degradation” provides a more holistic representation of how the disease reconfigures the aging brain.

### 2.2 Identifying and characterising disease subtypes with distinct disease progression trajectories

Building on our multimodal embedding and pseudotime analysis, we sought to identify subtypes that capture distinct trajectories of the inferred disease progression. Subtype discovery was performed exclusively among impaired subjects (including MCI and AD patients) using the multimodal imaging–derived disease space (see Methods 4.2), jointly informed by pseudotime trajectories, while CN individuals were assigned to a single reference subtype. The Bayesian information criterion (BIC) [20] was used to determine the optimal number of putative disease subtypes (Fig. 3a), resulting in four subtypes among impaired subjects, in addition to the CN reference group. All identified subtypes demonstrated significant stability (Methods 4.4.4), defined as the rate at which pairs of subjects were consistently assigned to the same subtype across repeated bootstrapped samples [37], with three subtypes showing strong significance (*p* < 0.001) and one subtype remaining significant at a moderate level (*p* < 0.05) based on permutation test.

**Fig 3.**
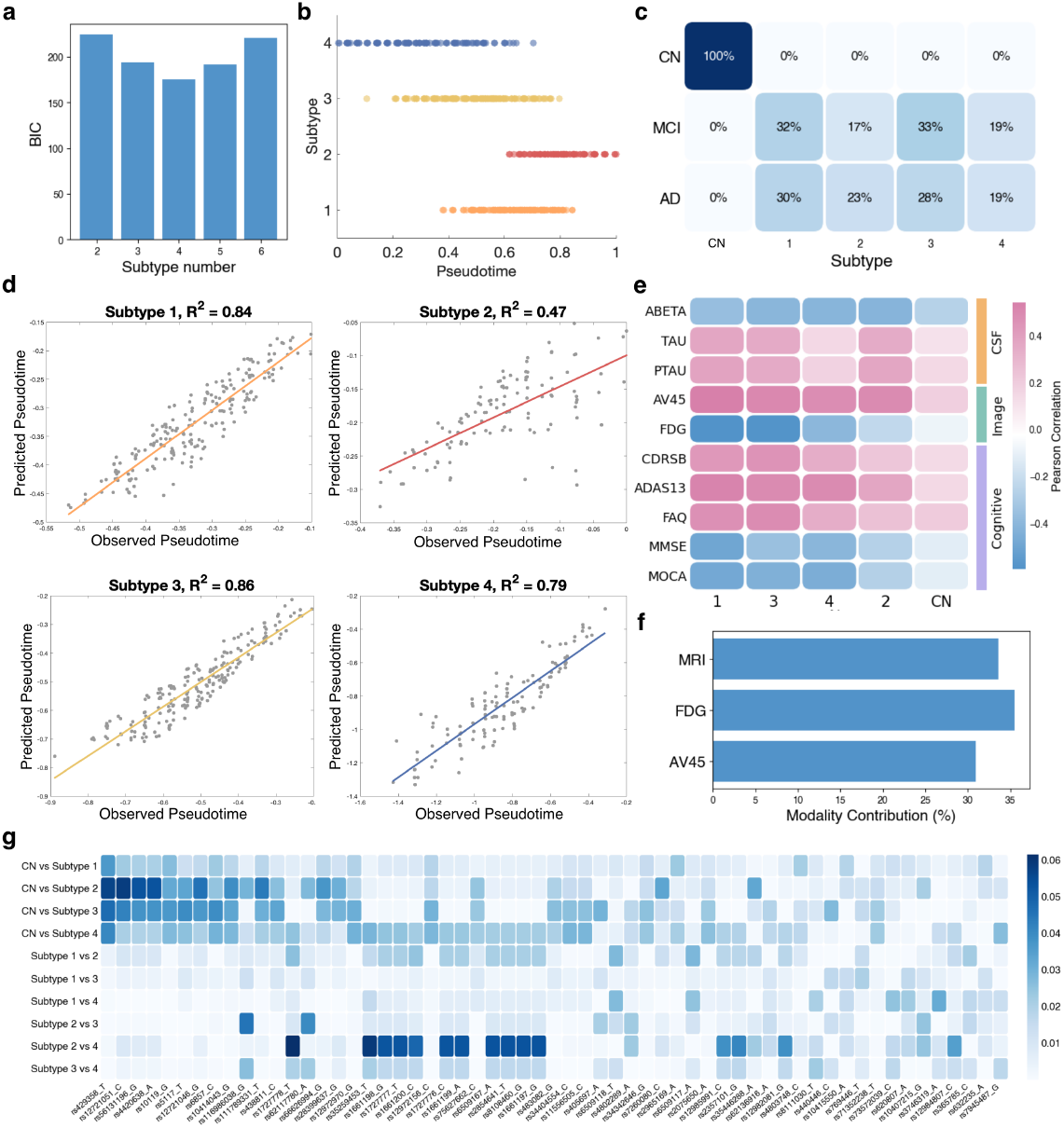
Identifying and characterising disease subtypes with distinct disease progression trajectories. (a) BIC values across different numbers of selected subtypes. The lowest BIC value indicates the optimal number of subtypes. (b) Distribution of pseudotime in each identified subtype, where each color represents one subtype. (c) Distribution of identified subtypes in each clinical diagnostic group. Numbers indicate the proportion of subjects per subtype within each diagnostic group. (d) Subtype-specific predictability of disease pseudotime. For each subtype, linear regression models were used to predict pseudotime from MDS coordinates, and the correlation coefficient (*R*^2^) quantifies the agreement between predicted and observed pseudotime. (e) Within-subtype correlations between biomarkers and pseudotime. Subtypes are in descending order based on FDG measures to facilitate comparison, with CN serving as the background group. (f) Percentage contribution of each imaging modality for subtype identification. (g) Effect sizes (*η*^2^) from pairwise ANOVA tests comparing subtypes based on SNPs. Only significantly associated features are shown (*p* < 0.05). ANOVA tests were adjusted for age, sex, and education level. The color scale represents the proportion of variance explained (*η*^2^).

Fig. 3b shows the continuous pseudotime trajectory of each disease subtype, where the pseudotime of each subject is represented by a dot. Each subtype occupies a partially overlapping yet distinct segment of the pseudotime continuum, highlighting the non-discrete nature of disease progression. Notably, Subtype 2 was predominantly concentrated at the advanced end of the pseudotime spectrum (0.6–1.0), representing a severe disease phenotype.

We then examined the distribution of identified subtypes in each clinical diagnostic group (Fig. 3c). Disease subtypes were distributed relatively evenly in each group of MCI and AD, indicating that clinically defined disease groups do not fully capture underlying biological heterogeneity. Together with pseudotime analysis, these findings suggest that the identified disease subtypes reflect distinct biological states along a continuous disease trajectory rather than discrete clinical stages.

To evaluate the internal homogeneity of the identified subtypes, we fitted a regression model of the observed pseudotime on the corresponding MDS coordinates of all subjects within each subtype (Methods 4.4.5). This allowed pseudotime of a subject to be predicted using linear regression, and the correlation coefficient (*R*^2^) between the predicted and observed pseudotime was calculated. The resulting *R*^2^ values (Fig. 3d) indicate good correspondence for Subtypes 1, 3, and 4 (*R*^2^ > 0.79), demonstrating these identified subtypes capture coherent progression patterns and enhance the interpretability of pseudotime. Notably, Subtype 2 exhibited a more modest fit (*R*^2^ = 0.47), which was accompanied by a marked spatial convergence in the MDS embedding (Fig. 2a), where subjects in this group clustered into a dense region compared to the more dispersed distribution of earlier-stage individuals.

We then evaluated the associations between subtypes and biomarkers. Fig. 3e shows subtype-specific correlations between pseudotime and multiple biomarkers. Across all disease subtypes, the direction of correlations was consistent with known biomarker trajectories along disease progression. Compared with the CN group used as a reference, disease subtypes exhibited overall stronger correlations, indicating a tighter coupling between pseudotime and biomarker changes. Importantly, each disease subtype displayed distinct associations with different biomarkers, indicating subtype-specific characteristics. Subtype 1 and 3 exhibited robust, correlations across nearly all biomarkers. In contrast, Subtype 2 revealed a notable dissociation between pathology and function: while it maintained a strong correlation with amyloid deposition (ABETA and AV45), its associations with FDG and clinical cognitive scores were significantly attenuated. Additionally, Subtype 4 exhibited a unique profile with markedly reduced correlations with CSF TAU and PTAU, potentially reflecting a tau-independent pathway of neurodegeneration that warrants further investigation.

To further provide biological support evidence of the derived subtypes, we quantified how strongly each Single Nucleotide Polymorphism (SNP) differentiates subtypes by pairwise effect sizes (*η*^2^) of SNPs with overall ANOVA screening (*p* < 0.05, see Methods 4.4.6). CN individuals served as a reference subtype for subsequent comparisons. As shown in Fig. 3g, while large genetic effects were observed when distinguishing CN from disease subtypes, we identified significant locus-specific variations between disease subtypes. Notably, Subtype 2 and 4 as well as Subtype 3 and 4 exhibited distinct clusters of high-effect-size SNPs, suggesting that their divergent progression trajectories may be rooted in unique genetic architectures. This highlights that the identified subtypes capture divergent pathological trajectories that may be driven by different underlying genetic predispositions.

To assess the relative contribution of each imaging modality to the definition of the identified subtypes, we applied a leave-one-modality-out approach (Methods 4.4.7). The normalized mutual information analysis (Fig. 3f) revealed a balanced contribution across modalities, with FDG PET (35.5%), structural MRI (33.6%), and AV45 PET (30.9%) each playing a substantial role. This confirms that our subtype identification is driven by the synergistic integration of metabolic, structural, and proteinopathy features rather than being dominated by a single pathological marker. Similar to SNP ANOVA screening, further evaluation of pairwise effect sizes for individual imaging-derived mode-based features (Fig.S3) was obtained. Within disease subtypes, effects were generally sparse and small. This indicates that subtype differentiation is emerging from the coordinated signature of multimodal features rather than being tied to isolated regional changes or independent features.

Taken together, these results demonstrate that subtype differentiation with distinct disease progression trajectories is strongly supported by clinical, biological, and cognitive evidences.

### 2.3 Geometric feature extraction outperforms conventional approaches in identifying disease progression and subtypes

To assess the effectiveness of mode-based features for pseudotime inference, we compared the mode-based pseudotime characteristics with those derived from conventional region- and vertex-based imaging features (see Methods 4.1.2). Specifically, region- based features consisted of measures from 68 regions of the Desikan-Killiany (DK) atlas [12] were extracted from each imaging modality, while vertex-based features were obtained using principal component analysis (PCA) with the number of components selected based on optimal dimensionality. The optimal numbers of vertex-based and mode-based features were decided based on the Spearman correlation between the pseudotime and clinical diagnosis, as per the analysis of mode-based features (see Methods 4.3 for the range of feature numbers and Table 1 for the optimal feature number K).

**Table 1.** Performance comparison of three imaging feature extractions in pseudotime and subtype identification. K indicates the feature dimension. Left: Associations between pseudotime and clinical diagnosis across three imaging feature extractions, including Spearman correlation (Corr) and permutation-corrected ANOVA p-values. Right: Subtype-level pseudotime prediction performance across three imaging feature extractions, quantified by the coefficient of determination (*R*^2^) for each disease subtype and the average *R*^2^.

| Feature | K | Pseudotime | | Subtype $R^2$ | | | | | Average |
| --- | --- | --- | --- | --- | --- | --- | --- | --- | --- |
|  |  | Corr | p-value | S1 | S2 | S3 | S4 | S5 |  |
| Region | 68 | 0.144 | 5.1e-11 | 39.560 | 48.404 | 95.727 | 95.038 | 17.052 | 59.156 |
| Vertex | 100 | 0.147 | 6.8e-05 | 52.767 | 65.364 | - | - | - | 59.065 |
| Mode | 30×2 | **0.255** | **9.5e-14** | 84.330 | 46.529 | 86.005 | 79.466 | - | **74.082** |

We first visualized the distribution of pseudotime in each clinical diagnosis with region-, vertex-, and mode-based features (Fig. 4a–c). Across all three feature extractions, pseudotime differed significantly between CN and AD subjects (permutation test, Bonferroni-corrected *p* < 0.001), with AD individuals exhibiting higher pseudotime values than both CN and MCI subjects. MCI subjects showed intermediate pseudotime values, consistent with a continuous disease progression trajectory. Notably, while region- and vertex-based features showed at least one pair of diagnostic groups without significant differences, mode-based features yielded significant pseudotime differences across all three diagnostic groups. As for the Spearman correlations between the pseudotime and diagnosis labels and ANOVA test for distinguishing clinical diagnostic by pseudotime (Methods 4.4.8), mode-based features present the highest association between the pseudotime and clinical diagnosis(Table 1).

**Fig 4.**
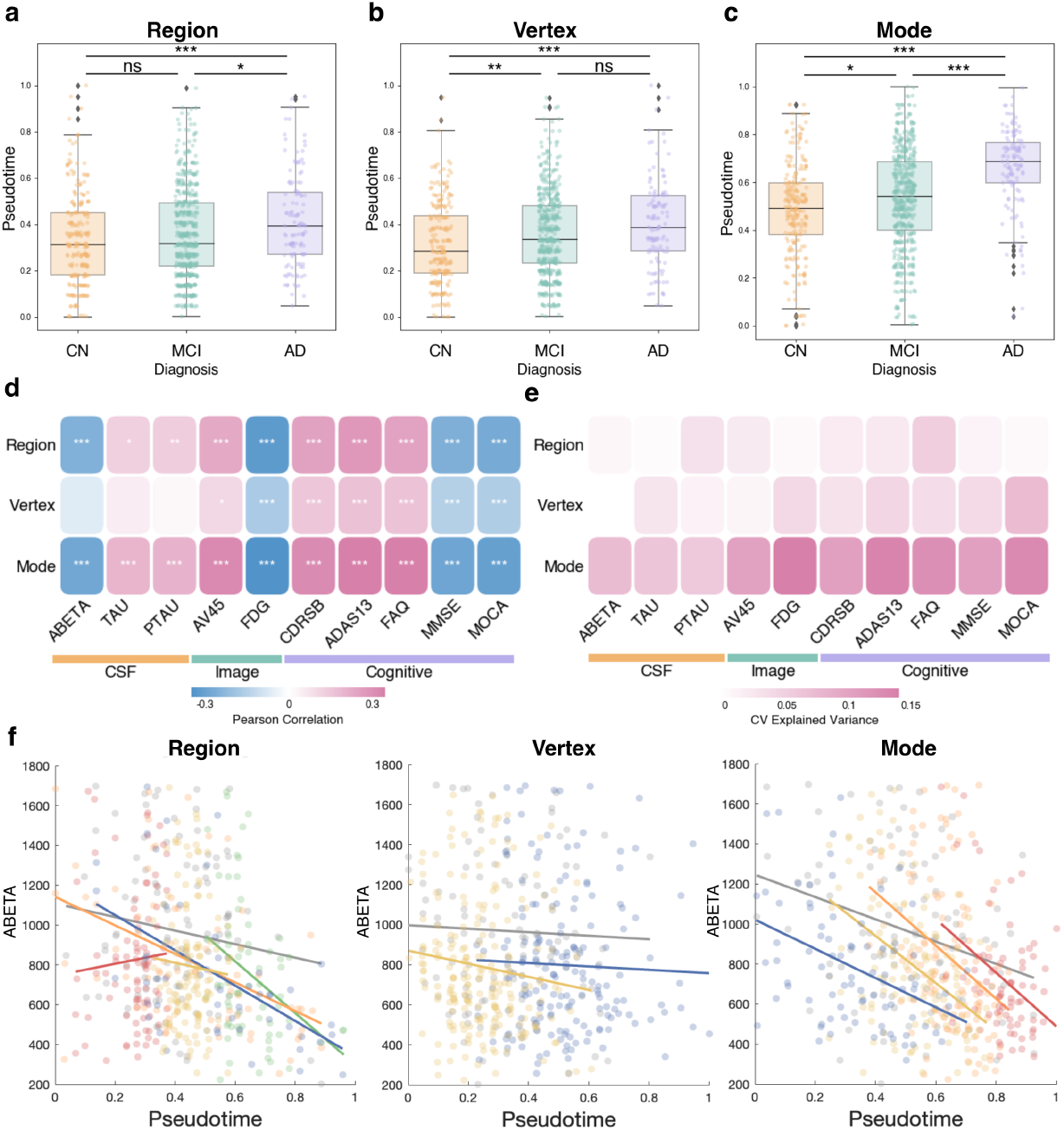
Identifying disease progression and subtypes using region-, vertex-, and mode-based imaging features. (a–c) Distribution of pseudotime in each clinical diagnosis with different feature extractions. Asterisks denote permutation-based p-values with Bonferroni correction (***: *p* < 0.001, **: *p* < 0.01, *: *p* < 0.05; ns: not significant). (d) Pearson correlations between biomarkers and pseudotime for different feature extraction methods. Colors indicate correlation coefficients, and asterisks denote Bonferroni-corrected p-values (***: *p* < 0.001, **: *p* < 0.01, *: *p* < 0.05). (e) Mean explained variance from 5-fold cross-validated regression models across different feature extraction methods, using pseudotime to predict individual biomarkers. (f) Amyloid,β levels along pseudotime overlaid with subtype labels. Lines represent within-subtype linear fits, with the gray line corresponding to the CN background group and colored lines representing individual disease subtypes.

We next examined the associations between the different pseudotime estimates and biomarkers (Fig. 4d). Overall, mode-based features showed the highest correlations across all biomarkers (| Pearson *ρ* |= 0.20–0.35), whereas region-based features showed intermediate correlations (| *ρ* |= 0.13–0.32) and vertex-based features showed the lowest (| *ρ* |= 0.02–0.17). Significant associations between cognitive scores and pseudotime were observed for all three feature extractions (Bonferroni-corrected *p* < 0.001). For CSF and imaging biomarkers, only mode-based features showed consistently strong correlations (Bonferroni-corrected *p* < 0.001). Specifically, FDG PET and amyloid-related measures (CSF ABETA and AV45 summary SUVR) showed higher correlations with pseudotime than tau-related features (CSF TAU and PTAU), consistent with the fact that input features were derived from amyloid and FDG PET data. Fig. 4e shows the cross-validated explained variance when using pseudotime to predict individual biomarkers. Among the three imaging feature extractions, mode-based features achieved the highest explained variance, indicating that the corresponding pseudotime exhibits more robust and generalizable associations with biological measurements. Together, these results indicate that mode-based features provide the most robust and sensitive pseudotime capturing both clinical diagnosis and biological mechanisms more effectively than region- or vertex-based features.

We next compared mode-, region-, and voxel-based features with respect to sub-type identification by evaluating R^2^ values (Table 1), which quantify the internal homogeneity of each identified subtype and are derived from the regression model as described in Section 2.2. Different feature extractions generated varying numbers of disease subtypes, with mode-based features achieving the highest average *R*^2^ (74.08), indicating high homogeneity of mode-based subtypes.

We overlaid subtype information of ten biomarkers versus pseudotime on scatter plots and illustrated the fit line for each subtype (see (Fig. 4e for ABETA and Supplementary Fig. S2 for other nine biomarkers). For region-based features, some generated subtypes showed inverse trends relative to the expected biological direction, and with many subtypes, slopes were highly variable. Vertex-based features produced correlations in the expected direction, but the distinction between healthy and disease subtypes was less clear. In contrast, disease subtypes derived from mode-based features exhibited relatively similar slopes for each biomarker, indicating that mode-based features capture a stable pattern of biomarker change along pseudotime in each subtype. These slopes were also clearly distinct from the CN subtype, facilitating differentiation between healthy and disease progression trajectories. Overall, mode-based features provide a more stable, distinctive, and interpretable subtyping structure compared with region-based and vertex-based features. These findings were replicated in an independent dataset, the Open Access Series of Imaging Studies 3 (OASIS-3) [29] (see Supplementary Analysis).

## 3 Discussion

AD patients suffer a significant delay in diagnosis and prognosis due to a lack of accessible biomarkers and diagnostic tools. In this work, we provided a framework to obtain a disease progression score––pseudotime––and the respective subtype of AD for individuals. We used the combination of eigenmode-based features extracted from three imaging modalities, AV45 PET, FDG PET, and sMRI, which measure brain phenotypes objectively and rather than relying on subjective cognitive measures. Our results showed that the pseudotime captures disease progression, with trajectories consistent with conventional clinical diagnosis and biomarker changes, and the provided sub-types display distinct disease phenotypes that are supported by imaging and genetic evidences.

The inferred pseudotime showed coherent associations with established CSF and imaging biomarkers. Strong correlations with amyloid PET, CSF amyloid *β*, and FDG PET are expected given their explicit inclusion in the multimodal imaging embedding. Notably, significant associations were also observed with CSF TAU and PTAU measures, despite the absence of tau-related imaging features in the model. One plausible explanation is that accelerated tau-related changes tend to occur after amyloid accumulation but prior, or in close temporal proximity, to the accelerated decline in cortical thickness [23, 34]. As a result, tau-related variation may be indirectly reflected in the inferred disease trajectory, even in the absence of explicit tau imaging features. Consistent with this interpretation, biomarkers that are known to change earlier than amyloid PET (e.g., CSF amyloid *β*), as well as downstream clinical measures reflecting cognitive impairment that typically emerges after substantial metabolic and structural decline [23], also exhibited significant associations with pseudotime. Together, these findings suggest that the proposed mode-based feature extraction, integrated within the mcTI algorithm, effectively capture biologically meaningful signals from each modality and facilitate the integration of complementary disease information.

The differentiation of disease subtypes in this study underscores the power of the mcTI algorithm in capturing the heterogeneous landscape of AD. Although imaging features contribute to the identification of the subtype as input, these features, when considered individually, mainly distinguish CN from disease subtypes, with relatively small differences between disease subtypes (Fig. S5). This likely reflects the mcTI algorithm’s ability to integrate features from different modalities. By projecting high-dimensional data into a low-dimensional latent space, our framework effectively integrates pathological, metabolic, and structural features, capturing nuanced patterns that traditional single-modality analyses might overlook.

Our results identify Subtype 2 as a terminal biological state characterized by phenotypic convergence and pathological-functional dissociation. The spatial clustering in the MDS embedding suggests that as AD reaches its end-stage, the brain’s initial heterogenous trajectories converge toward a stereotyped pathological terminal [48, 25], likely reflecting the exhaustion of individual compensatory mechanisms. In this saturated phase, the persistent correlation of amyloid and tau accumulations alongside weakened metabolic and cognitive associations indicates that functional indicators reach a symptomatic floor [23]. This suggests that while proteopathic burden may continue to accumulate, its linear coupling with cognitive performance is decoupled. Consequently, traditional cognitive scales lose their dynamic range, rendering them less sensitive for monitoring the terminal phase of the disease.

Analyzing differences between subtypes provides critical insights into potential biomarkers for disease progression. The biological validity of these subtypes is further reinforced by their distinct genetic and neuroimaging underpinnings. For example, among SNP features (Fig. 3g), we found that rs429358 and rs12721051 show significant differences across CN and multiple disease subtypes. Notably, rs429358 marks the *ε*4 allele of the apolipoprotein E (APOE) gene, a well-known genetic risk factor for AD progression [27, 6], while rs12721051 is located in the APOC1 gene and has been associated with AD-related genetic risk [19, 40]. In addition, several SNPs have not yet been extensively studied for their association with AD progression and may serve as potential novel biomarkers. For instance, we observed significant differences for rs1727778 and rs35259453 between subtype 2 and 4, and for rs116986038 and rs62117780 between subtype 2 and 3. These findings highlight candidate genetic variants that could contribute to subtype-specific pathological trajectories. Furthermore, our mode-based imaging feature analysis identified specific functional signatures that differentiate disease subtypes. The imaging features of mode 21 is the most distinct when comparing CN with each disease subtype (Fig. S5), which shows an overlap with the dorsal attention network known related to working and episodic memory and thus affected in AD [54, 31]. The convergence of these heterogeneous genetic and imaging markers underscores a robust biological tethering, suggesting that the identified subtypes are anchored in a consistent multi-modal pathological framework.

Despite these insights, a limitation of the current study lies in the treatment of CN subjects as a static reference group. As clinical normal individuals may later convert to MCI or AD, some CN subjects could potentially align with specific disease sub-types [59, 13, 58] that our current assignment step might underrepresent. Future work extending this framework to model the transition from CN to specific disease subtypes will be crucial for identifying the earliest windows for subtype-specific intervention.

Our novel way of extracting features from imaging data, based on eigenmodes, showed superior performance compared to conventional features such as region-based features and PCA of vertex-based maps. Across multiple analyses—including pseudotime–biomarker correlations, subtype homogeneity, and cross-cohort replication consistently outperformed conventional approaches in capturing AD progression. This advantage arises because eigenmodes describe dynamic patterns at multiple spatial scales rather than focusing on specific regions, allowing the detection of subtle and distributed changes that conventional approaches may miss. Moreover, compared with regional- and vertex-based features that are more susceptible to cohort-specific preprocessing, mode-based features tend to be more robust and reproducible across datasets due to the simple process of extracting features, which only depending on the cortical surface mesh. Mode-based features provide spatially interpretable patterns that can be linked to known neuroanatomical or functional networks but also have unique information. These patterns represent intrinsic dynamic responses (i.e., standing waves) constrained by brain geometry and thus may also influence disease spread. These properties suggest that eigenmode-based features could serve as reliable biomarkers for more early disease detection and disease trajectory identification analyses.

Cross-cohort replication demonstrated the generalisability of the model-based features combined with the mcTI algorithm, further supporting the robustness of the proposed framework. Nevertheless, there are some limitations, mainly due to the relatively limited sample size of disease subjects and differences in biomarker characteristics in the OASIS-3 cohort as detailed in the Supplementary analysis.

Taken together, our findings indicate that imaging geometric signatures, integrated within the mcTI algorithm, can capture AD progression and distinguish subtypes with meaningful biological and genetic associations. This approach provides a promising framework for understanding disease heterogeneity and lay a foundation for developing precise diagnostic and pronosis tools for AD.

## 4 Methods

### 4.1 Datasets

Data were obtained from two independent, publicly available cohorts: the Alzheimer’s Disease Neuroimaging Initiative (ADNI, https://adni.loni.usc.edu) [51, 52] and the Open Access Series of Imaging Studies 3 (OASIS-3, https://sites.wustl.edu/oasisbrains/home/oasis-3) [29]. Both cohorts include multimodal neuroimaging, demographic information, and clinical assessments spanning cognitively normal individuals and patients across the AD progression. Specifically, we included 243 CN, 514 MCI, and 124 AD subjects from ADNI cohort, as well as 374 CN and 77 AD subjects from OASIS-3 cohort after data quality control, see Table 2, S1 for detailed information.

**Table 2.** Demographic and biomarker characteristics of the ADNI dataset. Values are presented as mean ± standard deviation.

|  | CN | MCI | AD | Overall |
| --- | --- | --- | --- | --- |
|  | N=243 | N=514 | N=124 | N=881 |
| Sex (M/F) | 120/123 | 290/224 | 75/49 | 485/396 |
| Age (years) | 76.19 ± 6.50 | 72.82 ± 7.90 | 74.65 ± 8.57 | 74.01 ± 7.78 |
| Education (years) | 16.45 ± 2.65 | 16.13 ± 2.67 | 15.83 ± 2.57 | 16.17 ± 2.66 |
| Aβ (pg/mL) | 1,170.29 ± 455.24 | 952.94 ± 437.95 | 680.84 ± 328.58 | 954.97 ± 452.66 |
| Tau (pg/mL) | 257.73 ± 98.61 | 297.91 ± 143.17 | 387.74 ± 162.53 | 305.06 ± 144.13 |
| pTau (pg/mL) | 23.78 ± 10.58 | 28.47 ± 15.55 | 37.85 ± 16.85 | 29.09 ± 15.45 |
| AV45 (SUVR) | 1.12 ± 0.19 | 1.23 ± 0.23 | 1.38 ± 0.22 | 1.22 ± 0.23 |
| FDG (SUVR) | 1.27 ± 0.12 | 1.21 ± 0.16 | 1.05 ± 0.15 | 1.19 ± 0.17 |
| Whole brain volume (mm<sup>3</sup>) | 1,012,328 ± 106,152 | 1,036,227 ± 106,599 | 1,004,393 ± 114,377 | 1,025,114 ± 108,501 |
| CDRSB | 0.40 ± 1.33 | 2.43 ± 2.74 | 5.01 ± 2.45 | 2.24 ± 2.78 |
| ADAS13 | 10.20 ± 6.95 | 18.23 ± 12.24 | 32.74 ± 10.23 | 18.03 ± 12.74 |
| FAQ | 1.11 ± 3.52 | 5.60 ± 7.53 | 14.41 ± 7.58 | 5.61 ± 7.81 |
| MMSE | 28.58 ± 1.97 | 26.57 ± 4.11 | 22.23 ± 3.40 | 26.51 ± 4.04 |
| MOCA | 25.05 ± 3.16 | 22.21 ± 4.86 | 16.23 ± 5.25 | 22.17 ± 5.25 |

#### 4.1.1 Neuroimaging data

For ADNI, we included cross-sectional [^18^F]-Florbetapir (AV45) PET, [^18^F]-Fludeoxyglucose (FDG) PET, and structural MRI data and obtained preprocessed volumetric images from publicly available ADNI directory. T1-weighted structural MRI preprocessing algorithm included gradient nonlinearity correction (gradwarp) and intensity non-uniformity correction using the ADNI modified N3 algorithm (N3m) [5, 45]. Amyloid PET (AV45) and FDG PET preprocessing includes coregistration to corresponding sMRI, averaging across time frames, and intensity standardization, as provided by the ADNI PET core [26].

For OASIS-3, we included cross-sectional AV45 PET and sMRI data and obtained preproccessed volumetric data from the OASIS portal. Structural MRIs from OASIS-3 were processed using the FreeSurfer software, including motion correction, non-brain tissue removal, intensity normalization [29]. PET images were generated using the Pet Unified Pipeline (PUP) [29].

#### 4.1.2 Feature extraction

We extracted mode-based, region-based, and vertex-based features from surface-based maps derived from the preprocessed volumetric images of each individual. Vertexwise cortical thickness maps were obtained from the FreeSurfer recon-all pipeline [11, 16, 15], resampled to the FsAverage template. For PET data, preprocessed volumetric images were projected onto the cortical surface with values sampled at the mid-cortical depth (projection fraction = 0.5) and restricted to cortical vertices. Surface PET maps were resampled to the FsAverage template to ensure inter-subject correspondence. Each vertex-based maps contains 327,684 vertices and thus Principal Component Analysis (PCA) was applied to reduce the number of features. As commonly used in the literature, we extracted 68 region-based features using the Desikan–Killiany atlas by FreeSurfer parcellation pipeline [17, 12].

We obtained mode-based features by decomposing the vertex-level maps using the cortical geometric eigenmodes as a basis set. Geometric eigenmodes Ψ for each hemisphere were derived by solving the eigenvalue problem for the Laplace-Beltrami operator Δ on the FsAverage cortical surface mesh with 163842 vertices (also termed the Helmholtz equation) of each hemisphere

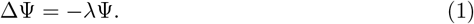

The Laplace–Beltrami operator, Δ, quantifies how much a measure at a given point differs from its surroundings. Each eigenmode, *ψ*_*j*_, represents a dynamic pattern of the cortical surface, and its corresponding eigenvalue *λ*_*j*_ indicates the spatial frequency or scale of that pattern, ordered from low frequency to high frequency scales. Low-frequency modes capture broad, global patterns on the brain surface while high-frequency modes capture finer, regionally localised details.

We used the basis set defined by these eigenmodes to perform a spectral decomposition of the PET- and MRI-derived brain maps using a general linear model,

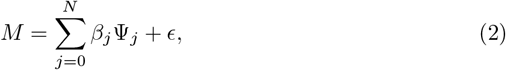

where *M* is the surface-based map, *N* is the number of eigenmodes used, *ψ*_*j*_ is the *j*-th eigenmode, *β*_*j*_ is its corresponding weight, representing the contribution of that mode to the observed map, and *ϵ* denotes the residual error. The coefficients, *β*_*j*_, quantify the contribution that each mode makes to reconstructing the observed spatial map.

#### 4.1.3 Demographic and biomarker data

Demographic and clinical variables for ADNI participants were obtained from the Quantitative Templates for the Progression of Alzheimer’s disease (QT-PAD) dataset (https://www.pi4cs.org/qt-pad-challenge), which were computed from an identical ADNI data freeze and subjected to qualitative comparison. We included age, sex, years of education, cognitive assessments, and cerebrospinal fluid (CSF) and imaging biomarker measurements. Specifically, we focused on AV45 and FDG PET SUVR as they reflect disease-related pathology, whereas whole brain volume mainly captures global anatomical scaling influenced by inter-subject variability such as head size. Cognitive measures available in ADNI included the Mini-Mental State Examination (MMSE), Clinical Dementia Rating Scale Sum of Boxes (CDRSB), Functional Activities Questionnaire (FAQ), Alzheimer’s Disease Assessment Scale–Cognitive Sub-scale (ADAS13), and the Montreal Cognitive Assessment (MoCA). CSF ABETA and CSF TAU measurements greater than 1700 or less than 8 were set to 1700 and 8, respectively.

We obtained whole-genome sequencing data for ADNI subjects and performed genome-wide SNP analysis. Variants had undergone read alignment, variant calling, and multi-stage quality control using a standardized pipeline based on the GRCh38 reference genome [30, 36]. We performed additional quality control by excluding variants with missingness >5%, minor allele frequency <1%, or Hardy–Weinberg equilibrium *p* < 1 × 10^*-*8^ in controls, as well as samples with genotype missingness >5%, duplicate subjects, or relatedness up to the second degree. Candidate Alzheimer’s disease–associated SNPs were prioritized using FinnGen GWAS summary statistics (*p* < 5 ×10^*-*8^) [28]. Missing genotypes were imputed using the most frequent allele. After filtering, 1,790 SNPs were retained for downstream analyses.

For the OASIS-3 cohort, demographic and cognitive data, including age, sex, MMSE, CDR, and FAQ, were obtained directly from the OASIS-3 database. Education level, additional cognitive assessments, and CSF or imaging biomarkers were not included for OASIS-3 due to substantial missingness or lack of consistent availability.

### 4.2 Multi-omics contrastive trajectory inference algorithms

We adapted the previously published mcTI algorithm [22], originally developed for multi-omics data, to multimodal imaging features. For each participant, the algorithm provides a pseudotime representing disease progression score and assigns the participant to putative disease subtypes. The detailed steps are:

- Data dimension reduction: Vertex-based imaging features were reduced via principal component analysis (PCA), whereas region-based and mode-based features were used directly without dimension reduction.
- Integration of multi-modal features: Imaging features from multiple modalities were integrated using similarity network fusion (SNF) [49] to construct a disease-relevant, patient-dominant fused similarity network (FN) that captures shared patterns of inter-subject relationships across modalities.
- Pseudotime calculation: Pseudotime, a disease progression score, was computed for all participants by measuring the shortest-path distance from the participant’s location on the fused similarity network to the cognitive normal (CN) subgroup’s centroid. These distances were subsequently standardized to a continuous scale between 0 and 1, yielding an interpretable measure of individual disease progression. Higher pseudotime value indicates more advanced stage to AD.
- Subtype assignment: Distinct patient subtypes were identified using an expectation–maximization (EM) procedure based on the FN. First, spectral clustering was applied to FN to obtain an initial subtype assignment. The FN was then embedded into a low-dimensional Cartesian space using multidimensional scaling (MDS), yielding a coordinate representation (FN-MDS). The optimal number of subtypes was determined by minimizing the Bayesian Information Criterion (BIC) [20]. The EM procedure was subsequently performed in the FN-MDS space to refine subtype assignments. For each subtype, a predictive model was trained to estimate the pseudotime based on the FN-MDS coordinate values. Using a leave-one-out strategy, each subject was evaluated across all subtype-specific models, resulting in prediction errors in pseudotime that quantify how well the subject fits each subtype. Subjects were reassigned to the subtype yielding the lowest prediction error, and subtype-specific models were retrained based on the updated assignments. This process was repeated until convergence.

Our adaptations preserve the conceptual structure of mcTI while applying it to multimodality imaging features instead of molecular features.

### 4.3 Optimising number of features

For region-based features, measurements from all 68 cortical regions were included, whereas for vertex-based and mode-based features, the features were represented using an optimally selected dimensionality. Mode-based features were derived independently for each hemisphere and subsequently combined across hemispheres, with feature dimensionalities of 10, 20, 30, 40, 50, and 100 per hemisphere evaluated. Correspondingly, for vertex-based features, whole-brain representations with dimensionalities of 20, 40, 60, 80, 100, and 200 were examined. For each candidate dimensionality, pseudotime values were computed and their Spearman correlation with subjects’ diagnostic labels was evaluated, allowing selection of the dimensionality that best captures disease aligning with clinical diagnosis. Statistical significance of the observed correlations was assessed independently using permutation testing (See Methods 4.4.8 in detail), mitigating potential bias from selecting the dimensionality that maximizes the correlation. The final dimensionality chosen for each feature type is summarized in Tables 1 and S2.

### 4.4 Validating and characterising the disease progression score and subtypes

#### 4.4.1 Differences in pseudotime across diagnostic groups

Differences in pseudotime across diagnostic groups were assessed to determine whether the inferred disease progression scores captured clinically meaningful distinctions. Pairwise comparisons of pseudotime between diagnostic groups (CN, MCI, and AD) were performed using nonparametric permutation tests. Diagnostic labels were randomly permuted 10,000 times to generate an empirical null distribution, against which the observed statistic was evaluated. Two-sided permutation-based p-values were calculated as the proportion of permutations yielding an absolute mean difference greater than or equal to the observed value. To control for family-wise error arising from multiple pairwise group comparisons, permutation-based p-values were further adjusted using Bonferroni correction.

#### 4.4.2 Pearson correlation between pseudotime and biomarkers

Associations between pseudotime and continuous cognitive, CSF, or imaging-derived biomarkers without ordinal information were quantified using Pearson correlation coefficients. Statistical significance was assessed using analytical p-values, followed by Bonferroni correction to control for family-wise error across biomarkers.

#### 4.4.3 Cross-validation regression for predicting biomarkers from pseudotime

To further assess the robustness and generalizability of the relationship between pseudotime and biomarkers, we performed 5-fold cross-validated regression. For each biomarker, we treated the pseudotime as the independent variable **X**, the biomarker measurement as the dependent variable **Y**, and included age, sex, and education level as covariates. The explained variance 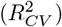 was calculated by:

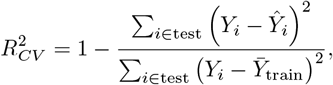

where *Y*_*i*_ is the observed biomarker value in the test set, *Ŷi* is the predicted value from the regression model, and 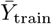 is the mean biomarker value in the training folds. The mean and standard deviation of explained variance values across folds were reported for each biomarker.

#### 4.4.4 Stability and significance of disease subtypes

To assess the stability of each identified subtype, we performed 500 bootstrap resampling and 5,000 permutation tests. A robust algorithm is expected to assign subjects consistently to the same subtype across bootstrap resampling, and these assignments should be more stable than expected under random permutation. For each pair of subjects originally assigned to the same subtype, pairwise stability [37] was quantified as the proportion of bootstrap iterations in which both subjects were included in the sample and assigned to the same subtype. The observed stability was then calculated as the average of these pairwise stability values across all pairs of subjects within the same subtype.

To assess the significance of subtype stability, we generated a null distribution by randomly permuting the original subtype assignments of patient subjects. For each permutation, null pairwise stability was calculated based on these shuffed subtype assignments. The empirical *p*-value for each subtype was then computed as the fraction of permutations in which the observed stability exceeded the null stability, reflecting the robustness of the clustering assignment.

#### 4.4.5 Homogeneity of pseudotime in each subtype

After the subtype assignments, to evaluate the internal consistency of each identified subtype, we performed linear regression within each subtype using the subjects’ MDS coordinates as predictors and the corresponding pseudotime as the response. For each subtype, we fitted the model and computed the Pearson correlation (*R*^2^) between the predicted and observed pseudotime. Higher *R*^2^ indicates greater internal homogeneity of the subtype.

#### 4.4.6 Associations between SNPs and disease subtypes

The associations between single nucleotide polymorphisms (SNPs) and disease sub-types were analysed using one-way ANOVA after adjusting for age, sex, and education. Effect sizes were quantified using *η*^2^, defined as the proportion of the total variance in each SNP that is attributable to differences between subtypes. Statistical significance of the associations was assessed using FDR-corrected *p*-values (Benjamini–Hochberg procedure). However, given the high dimensionality of genetic features, no associations survived multiple-comparison correction, and corrected results of SNPs are therefore not reported.

#### 4.4.7 Modality contribution

To evaluate the contribution of each imaging feature modality to subtype assignment, we performed a leave-one-modality-out analysis. For each modality *i*, subtype assignments were recalculated without that modality, and the normalized mutual information nMI between the original assignments and the leave-one-out assignments was computed. The contribution of modality *i* was quantified as 1 nMI, with higher values indicating a greater impact on the resulting subtypes. Contributions were further normalized across modalities for comparison.

#### 4.4.8 Spearman correlation between pseudotime and diagnosis

To assess the overall association between pseudotime and clinical diagnostic labels, Spearman rank correlation was computed between pseudotime and diagnosis encoded as an ordinal variable to capture monotonic progression effects. To further evaluate group-level differences in pseudotime across diagnostic categories, a one-way ANOVA was conducted, and significance was determined using a permutation-based framework to avoid parametric assumptions, where diagnostic labels were randomly permuted 10,000 times, and the F-statistic was recomputed for each permutation to generate a null distribution. The empirical p-value was calculated as the proportion of permuted F-statistics exceeding the observed F-statistic.

## Data Availability

The datasets analyzed in this study are available from the Alzheimers Disease Neuroimaging Initiative (ADNI) and the Open Access Series of Imaging Studies (OASIS-3) upon application and approval.

https://adni.loni.usc.edu/

https://sites.wustl.edu/oasisbrains/home/oasis-3/

## Acknowledgements

This work was supported in part by the NIH grants U01 AG068057, U01 AG066833, R01 AG071470, U19 AG074879, R01 LM013463, and P30 AG073105. The ADNI data sets were obtained from the Alzheimer’s Disease Neuroimaging Initiative database (https://adni.loni.usc.edu). The OASIS-3 data sets were obtained from the Open Access Series of Imaging Studies (https://sites.wustl.edu/oasisbrains/home/oasis-3).

Data collection and sharing for this project were funded by the Alzheimer’s Disease Neuroimaging Initiative (ADNI) (National Institutes of Health Grant U01 AG024904) and DOD ADNI (Department of Defense award number W81XWH-12-2-0012). ADNI is funded by the National Institute on Aging, the National Institute of Biomedical Imaging and Bioengineering, and through generous contributions from the following: AbbVie, Alzheimer’s Association; Alzheimer’s Drug Discovery Foundation; Araclon Biotech; BioClinica, Inc.; Biogen; Bristol-Myers Squibb Company; CereSpir, Inc.; Cogstate; Eisai Inc.; Elan Pharmaceuticals, Inc.; Eli Lilly and Company; EuroImmun; F. Hoffmann-La Roche Ltd and its affiliated company Genentech, Inc.; Fujirebio; GE Healthcare; IXICO Ltd.; Janssen Alzheimer Immunotherapy Research & Development, LLC.; Johnson & Johnson Pharmaceutical Research & Development LLC.; Lumosity; Lundbeck; Merck & Co., Inc.; Meso Scale Diagnostics, LLC.; NeuroRx Research; Neurotrack Technologies; Novartis Pharmaceuticals Corporation; Pfizer Inc.; Piramal Imaging; Servier; Takeda Pharmaceutical Company; and Transition Therapeutics. The Canadian Institutes of Health Research is providing funds to support ADNI clinical sites in Canada. Private sector contributions are facilitated by the Foundation for the National Institutes of Health (www.fnih.org). The grantee organization is the Northern California Institute for Research and Education, and the study is coordinated by the Alzheimer’s Therapeutic Research Institute at the University of Southern California. ADNI data are disseminated by the Laboratory for Neuro Imaging at the University of Southern California.

