## Supplementary Information for "Geometric brain signatures of Alzheimer’s disease progression and subtypes"

### S1 Supplementary Analysis on cross-cohort replication and transferability

To evaluate the robustness and generalizability of our findings, we replicated our analyses in an independent cohort, OASIS-3, which included 451 participants (374 CN and 77 AD subjects; see Methods 4.1 for details). Note that, due to the limited availability of FDG PET scans in OASIS-3 AD participants, the subsequent analyses for both ADNI and OASIS-3 cohorts were restricted to AV45 PET and sMRI. We aimed to determine whether the superior performance of mode-based features and the associations between pseudotime and biological markers observed in ADNI were preserved in OASIS-3.

#### S1.1 Replication of feature performance across cohorts

Following the procedure described in Section 2.3, we extracted three types of imaging features from AV45 PET and MRI scans for each cohort: 1) 68 region-based features using the DK atlas, 2) vertex-based features obtained via PCA with an optimal number of components (ADNI: 60 components; OASIS-3: 50 components), and 3) mode-based features with 10 dimensions per hemisphere. For vertex-based and mode-based features, the optimal dimensionality was determined based on the Spearman correlation between pseudotime and clinical diagnosis. For each cohort and feature type, the mcTI algorithm was independently applied to derive pseudotime and subtypes, which were then compared across feature types. Mode-based features yielded the highest and statistically significant Spearman correlations between pseudotime and diagnosis in both cohorts (Table S2, permutation FWE-corrected $p < 1 \times 10^{-4}$), as well as the best internal homogeneity within the subtypes in terms of $R^2$ (Table S3-S4).

We further evaluated the associations between pseudotime and cognitive biomarkers. Due to limitations in OASIS-3, only three cognitive scores (CDR, MMSE, and FAQ) were considered, and all analyses were adjusted for age and sex because of substantial missingness in education data. As shown in Fig. S3a-b, mode-based features consistently produced the highest correlations and regression explained variance values among the three feature extractions in both cohorts. The associations were highly significant in ADNI (FDR-corrected $p < 0.001$) and significant in OASIS-3 (FDR-corrected $p < 0.05$). These results demonstrate that mode-based features maintain their efficiency in capturing pseudotime and disease subtyping patterns even across independent cohorts.

#### S1.2 Clinical and biomarker consistency

To further assess cross-cohort consistency, we removed MCI subjects from the ADNI dataset, harmonized the diagnostic labels to match those in the OASIS-3 cohort, and re-ran the mcTI algorithm on the resulting two-class ADNI data.

We first visualized the MDS embeddings of the fused networks derived from AV45 PET and sMRI mode-based features for each cohort, with pseudotime overlaid (Fig. S3c). While region-based and vertex-level features showed limited cross-cohort alignment in MDS embeddings, mode-based features exhibited highly consistent embedding geometry and pseudotime structure across cohorts. Notably, these patterns were also consistent with those obtained from the three-modality ADNI analysis described in Section 2.1, indicating robust trajectory consistency across cohorts and across different numbers of imaging modalities.

We then updated the correlation analysis between pseudotime and cognitive scores (CDR, MMSE, and FAQ) using the two-class ADNI dataset, adjusting for age and sex. Consistent with the results reported in Section S1.1, significant associations were observed in both cohorts, with Bonferroni-corrected $p < 0.001$ in ADNI and $p < 0.05$ in OASIS-3. Importantly, the directions of the correlations were consistent across cohorts, with broadly comparable effect sizes (Fig. S4a), demonstrating concordant cognitive–pseudotime relationships.

In addition, we assessed correlations between individual imaging mode-based features and pseudotime and summarized the results using heatmaps (Fig. S3d). The two cohorts exhibited broadly consistent correlation patterns across imaging features, with AV45-related features showing particularly strong and reproducible associations with pseudotime in both datasets.

Taken together, these results demonstrate that the inferred pseudotime generated from mode-based features exhibits stable continuity and biological plausibility across independent cohorts, diagnostic groupings, and multimodal imaging features, supporting a monotonic relationship in which higher pseudotime corresponds to more severe AD–related impairment.

There are some limitations, mainly due to the relatively limited sample size of disease subjects and differences in biomarker characteristics in the OASIS-3 cohort. For instance, when evaluating cross-cohort consistency between cognitive biomarkers and pseudotime, correlations in OASIS-3 were generally lower than in ADNI. Supplementary Fig. S3c shows within-diagnosis correlations, where correlations in OASIS-3—particularly in the AD group—were notably smaller. Examining the distributions of biomarker values within each group (Fig. S4b), we found that the CN group in OASIS-3 had a narrower range of cognitive scores compared to ADNI, constraining the achievable correlation. Conversely, the AD group in OASIS-3 had a wider distribution but a relatively small sample size, which may also contribute to lower within-group correlations. Together, these factors likely led to the reduced overall correlations observed in OASIS-3. Additionally, the low proportion of AD subjects in OASIS-3 resulted in the mcTI algorithm identifying only two optimal subtypes, indicating that disease groups were not well separated. Future studies including more people with AD would enable a more robust evaluation of cross-cohort consistency in subtype identification.

### S2 Supplementary tables

**Table S1** Demographic and biomarker characteristics of the OASIS-3 dataset. Values are presented as mean ± standard deviation.

| | CN | AD | Overall |
|---|---|---|---|
| | N=374 | N=77 | N=451 |
| Sex (M/F) | 133/241 | 26/51 | 159/292 |
| Age (years) | 70.34 ± 8.22 | 75.73 ± 5.98 | 71.26 ± 8.13 |
| Education (years) | 16.11 ± 2.42 | 15.86 ± 2.08 | 16.06 ± 2.35 |
| CDRSB | 0.05 ± 0.31 | 2.49 ± 2.46 | 0.46 ± 1.39 |
| FAQ | 0.15 ± 0.70 | 6.09 ± 7.32 | 1.16 ± 3.80 |
| MMSE | 29.16 ± 1.49 | 26.32 ± 3.63 | 28.68 ± 2.28 |

**Table S2** Summary of pseudotime–diagnosis associations across imaging feature types in the ADNI and OASIS-3 cohorts, including feature dimensionality, Spearman correlation, and permutation-corrected ANOVA p-values.

| | ADNI | | | OASIS-3 | | |
|---|---|---|---|---|---|---|
| Feature | K | Corr | p-value | K | Corr | p-value |
| Region | 68 | 0.080 | 2.8e-02 | 68 | 0.183 | 5.8e-04 |
| Vertex | 60 | 0.113 | 2.1e-02 | 50 | 0.115 | 2.7e-02 |
| Mode | 10×2 | **0.177** | **1.5e-08** | 10×2 | **0.266** | **7.6e-09** |

**Table S3** Subtype-level pseudotime prediction performance in the ADNI cohort using AV45 and MRI features, quantified by the coefficient of determination ($R^2$) for the entire cohort before subtyping (Non), for each disease subtype (S1-S6), and the average across subtypes.

| Feature | Non | S1 | S2 | S3 | S4 | S5 | S6 | Average |
|---|---|---|---|---|---|---|---|---|
| Region | 3.703 | 93.794 | 24.894 | 94.318 | 2.132 | 24.544 | 59.399 | 49.847 |
| Vertex | 21.793 | 46.809 | 62.633 | 53.637 | 93.565 | - | - | 64.161 |
| Mode | 18.602 | 90.090 | 93.978 | 86.818 | 76.575 | 31.315 | - | **75.755** |

**Table S4** Subtype-level pseudotime prediction performance in the OASIS-3 cohort using AV45 and MRI features, quantified by the coefficient of determination ($R^2$) for the entire cohort before subtyping (Non), for each disease subtype (S1-S2), and the average across subtypes.

| Feature | Non | S1 | S2 | Average |
|---|---|---|---|---|
| Region | 10.318 | 48.200 | 57.208 | 52.704 |
| Vertex | 4.285 | 12.397 | 63.059 | 37.728 |
| Mode | 16.985 | 72.949 | 53.533 | **63.241** |

### S3 Supplementary figures

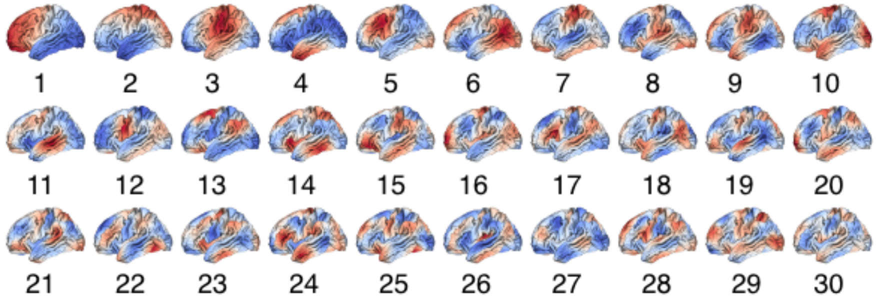


**Fig. S1** 30 eigenmodes (mode 1 to 30) of the left hemisphere derived from FsAverage surface mesh, ordered from low-frequency long-wavelength to high-frequency short-wavelength patterns.

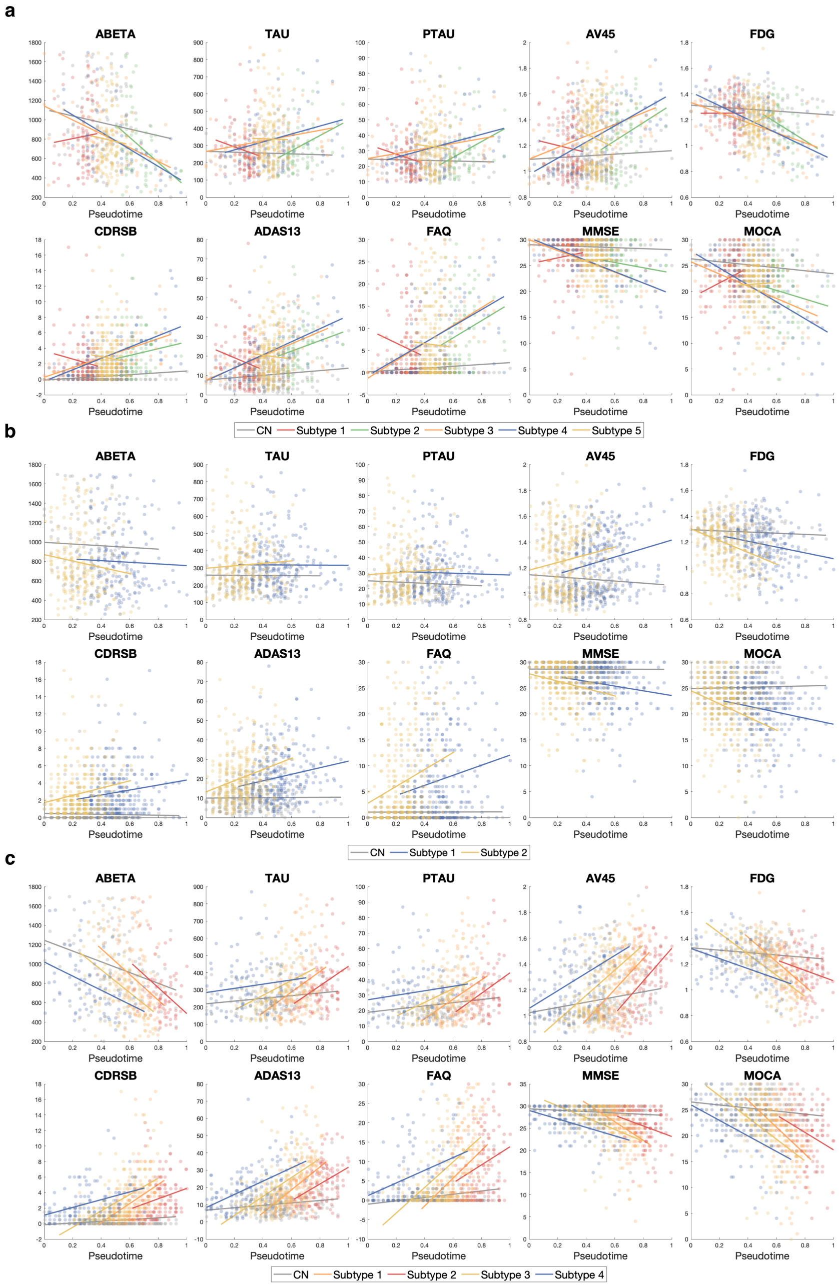


**Fig. S2** Biomarkers along pseudotime overlaid with subtype information for (a) region- (b) vertex- (c) mode-based features. Lines represent within-subtype linear fits, with the gray line corresponding to the CN background group and colored lines representing individual disease subtypes.

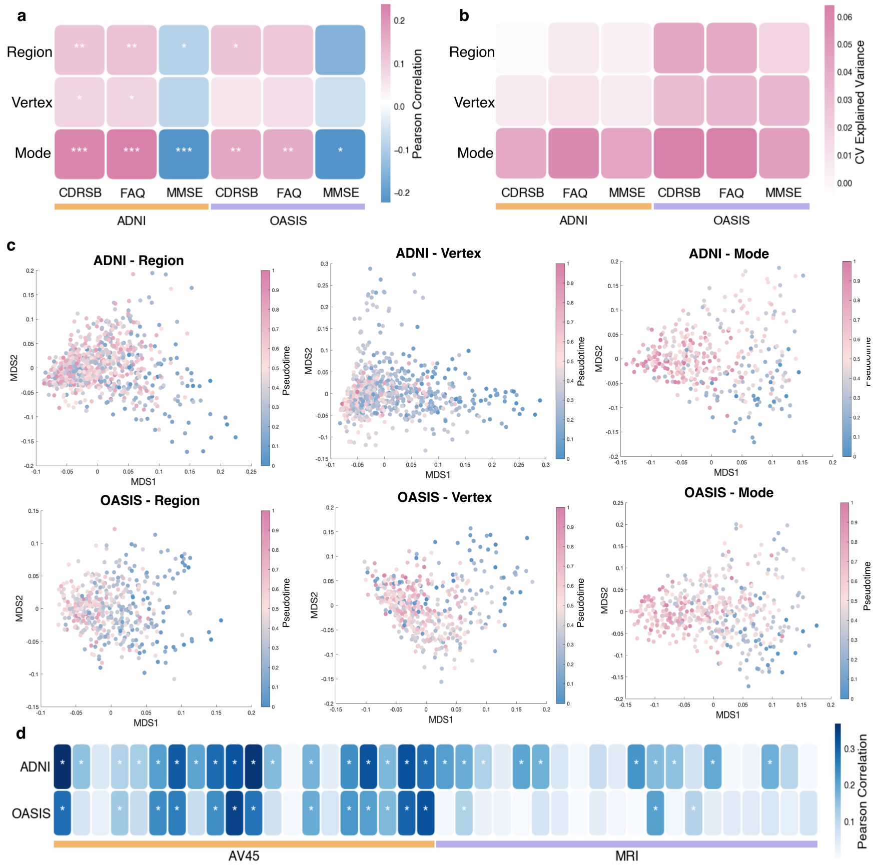


**Fig. S3** (a) Correlations between three cognitive biomarkers and pseudotime for each feature type. Colors indicate correlation coefficients and asterisks denote Bonferroni-corrected p-values (***: $p < 0.001$, **: $p < 0.01$, *: $p < 0.05$). (b) Mean explained variance from cross-validated regression models across different feature extraction methods, using pseudotime to predict individual biomarkers. (c) Two-dimensional MDS embeddings of the fused multimodal imaging feature networks based on region-, vertex-based, and mode-based representations for the ADNI and OASIS-3 cohorts, respectively. (d) Comparison of correlations between pseudotime and imaging mode-based features across cohorts. Results are shown for all imaging features, including 10 modes per hemisphere for two modalities. Darker color indicates the higher correlation coefficient and asterisks indicate statistically significant associations ($p < 0.05$, Bonferroni-corrected)

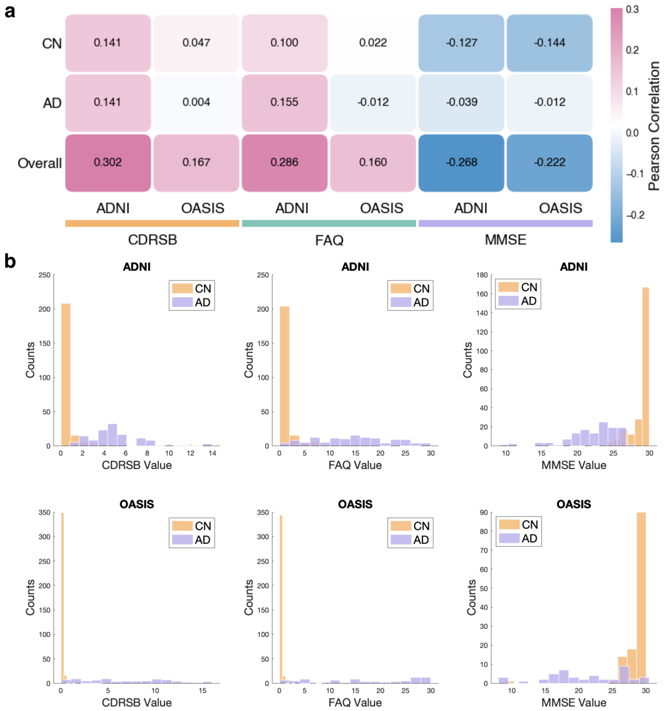


**Fig. S4** (a) Correlations between cognitive biomarkers and pseudotime within diagnosis groups and in the overall sample for each data cohort. Colors represent correlation coefficients. (b) Distribution of cognitive biomarkers and pseudotime across diagnosis groups in the ADNI and OASIS-3 cohorts.

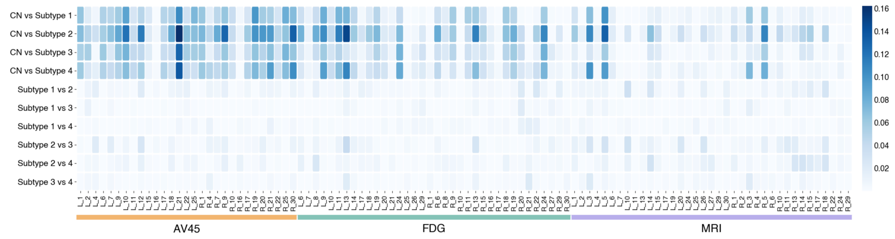


**Fig. S5** Pairwise subtype differences in imaging features and genetic variants. Effect sizes ($\eta^2$) from pairwise ANOVA tests comparing subtypes based on imaging features. Only significantly associated features are shown ($p < 0.001$, FDR-corrected). ANOVA tests were adjusted for age, sex, and education level. The color scale represents the proportion of variance explained ($\eta^2$).
